# Feasibility of using automatically extracted routine clinical data in a respiratory cohort study: The SPHN–SPAC demonstrator project

**DOI:** 10.64898/2026.07.14.26357927

**Authors:** Franco Romero, Mari Sasaki, Maria Christina Mallet, Eva Sophie Lunde Pedersen, Lorenz M. Leuenberger, Ronny Makhoul, Xenia Bovermann, Anna Hartung, Philipp Latzin, Seraphina Kissling, Alexander Moeller, Angela Treis, Nicolas Regamey, Fabiën N. Belle, Claudia E. Kuehni

## Abstract

**Objectives:** To assess the feasibility of using clinical data automatically extracted via the Swiss Personalized Health Network (SPHN) to complement or replace manually abstracted clinical data in the Swiss Paediatric Airway Cohort (SPAC).

**Materials and Methods:** We studied 1,075 SPAC participants enrolled between 2017**–**2023 at two Swiss children’s hospitals. Clinical data were extracted from electronic health records via SPHN in Resource Description Framework format, transformed into visit-centered datasets, and compared with manually abstracted SPAC clinical data and parent-reported emergency department (ED) visits and hospitalizations from follow-up questionnaires. We assessed feasibility by identifying challenges in acquiring data and evaluated data quantity, completeness, and agreement between datasets.

**Results:** We obtained analysis-ready SPHN-derived datasets from two hospitals after 24 months. SPHN-derived data captured more pneumology outpatient visits than manual abstraction (Hospital A: 1,963 vs 1,049; Hospital B: 2,343 vs 1,010) and identified clinical events among children without follow-up questionnaires. Completeness of variables varied across hospitals and encounters, reflecting differences in local clinical documentation practices. SPHN-derived and manually abstracted data showed high agreement for structured clinical variables, including spirometry measurements (concordance correlation coefficient >0.99). Self-reported and SPHN-derived ED visits and hospitalizations showed high absolute agreement but moderate concordance.

**Discussion and Conclusion:** Automated extraction of routine clinical data increased the completeness of longitudinal information compared with manual abstraction, suggesting that SPHN-derived data can complement manual data collection in cohort studies. Broader use remains limited by heterogeneous clinical documentation practices and the substantial effort required to harmonize and transform extracted data into analysis-ready research datasets.

## INTRODUCTION

Automated extraction of routine clinical data may streamline research workflows and reduce the burden of manual data collection in clinical studies, which often requires substantial resources and time (1,2). Large initiatives such as the Pediatric Learning Health System (PEDSnet) and the Patient-Centered Outcomes Research Network (PCORnet) in the United States have demonstrated that automatically extracted routine clinical data from electronic health records (EHRs) can support high-quality research while reducing manual workload (3–5). However, the quality of automatically extracted data depends on how information is documented within hospital systems and on the technical systems used for data extraction (6–8).

In Switzerland, the Swiss Personalized Health Network (SPHN) is a federal initiative that aims to facilitate the use of routine clinical data for research by applying the FAIR (Findable, Accessible, Interoperable, and Reusable) principles and establishing shared national data standards (9–12). Within this framework, initiatives such as SwissPedHealth implement the SwissPedData data model across Swiss children’s hospitals, enabling harmonized paediatric clinical data for research, quality improvement, and personalized care (13,14). However, the extent to which SPHN-derived data can complement and reduce reliance on manual data collection in clinical studies has not yet been systematically assessed. The SPHN-SPAC Demonstrator Project used the Swiss Paediatric Airway Cohort (SPAC) as a test case to determine the added value of SPHN’s data resources and infrastructure (15). In this study, we assessed the feasibility of using automatically extracted SPHN-derived data in a paediatric research setting. Specifically, we **(i)** evaluated the time required and practical challenges involved in acquiring analysis-ready datasets and **(ii)** compared the quantity, completeness, and accuracy of SPHN-derived information with manually abstracted and self-reported SPAC data.

## METHODS

The Swiss Paediatric Airway Cohort (SPAC) enrolls children aged 0–17 years who are referred to paediatric pneumology outpatient clinics for common respiratory symptoms across ten clinical centres in Switzerland. Trained study staff manually extract and abstract clinical information from electronic medical records at the pneumology outpatient visit that led to enrolment into SPAC (SPAC enrolment visit) and at least one subsequent pneumology follow-up visit. Information was retrieved from routine clinical documentation within hospital information systems, including clinical letters summarizing the outpatient consultation, as well as lung function and laboratory reports, and was manually transcribed into the study database. In addition, at baseline and annually thereafter, parents complete standardized questionnaires covering demographics, respiratory symptoms, comorbidities, environmental exposures, and respiratory-related clinical visits during the preceding 12 months. All SPAC data are stored in REDCap hosted by the Clinical Trials Unit at the University of Bern (16,17).

### SPHN-SPAC demonstrator project

For the SPHN–SPAC demonstrator project, we included SPAC participants enrolled between 1 July 2017 and 31 December 2023 at three participating centres (Bern, Zurich, and Lucerne). We selected these centres because they were the largest recruiting sites and had the technical infrastructure required to support automated data extraction within the Swiss Personalized Health Network (SPHN) framework. **Figure 1** shows the flow of data for the SPHN-SPAC study. For all included participants, we aimed to obtain and analyze three distinct datasets: **(i)** SPHN-derived data, consisting of routine clinical data for pneumology outpatient visits, respiratory-related emergency department (ED) visits, and respiratory-related hospitalizations, extracted from hospital electronic health record (EHR) systems using nationally standardized SPHN semantic data models (18); **(ii)** a restricted set of manually abstracted SPAC clinical variables corresponding to the same time window; and **(iii)** self-reported respiratory-related ED visits and hospitalizations occurring after the SPAC baseline visit, collected through annual SPAC follow-up questionnaires. We performed all data processing and analyses within the secure BioMedIT computing infrastructure, in accordance with SPHN data protection and governance requirements (19).

**Figure 1.**
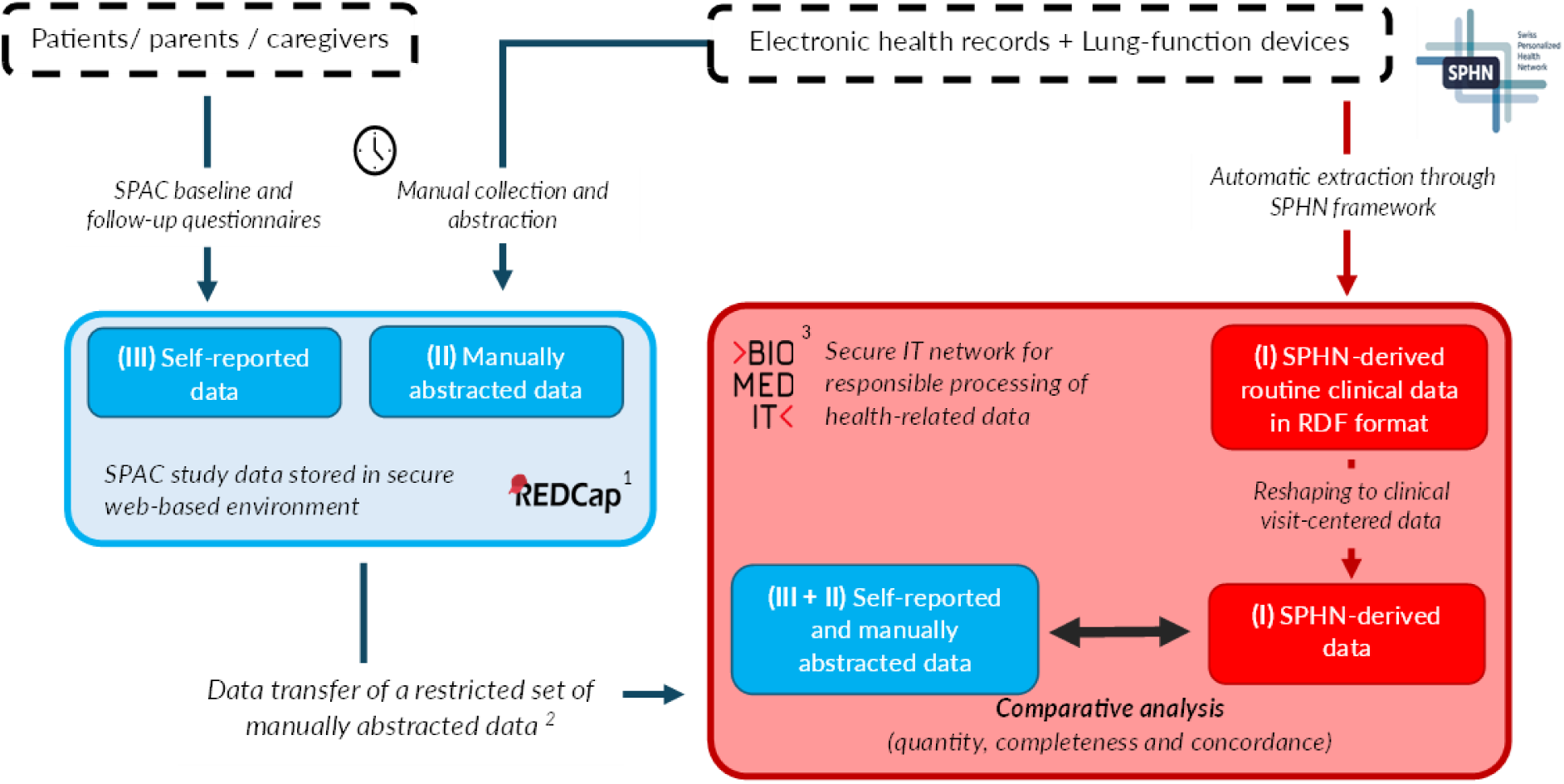
SPHN–SPAC dataflow schema.

Prior to SPHN-derived data extraction, we obtained the required ethics approvals and data-sharing agreements with each participating center. The step-by-step process for requesting extraction and preparation of SPHN-derived data for analysis is described in **supplementary Box 1**. As the first step, we defined the extractable variables based on the SPHN concept catalogue and local EHR structures in close collaboration with clinicians, hospital IT teams and SPHN data engineers (20,21). In the second step, hospital IT teams mapped local EHR fields to SPHN terminology standards, followed by the formal submission of data extraction requests through the SPHN infrastructure. In the third step, data were exported via the SPHN platform in Resource Description Framework (RDF) format, a standardized, machine-readable data format in which clinical information is stored as structured concepts (such as patients, visits, diagnoses, and measurements) together with explicit links between them, and securely transferred to the BioMedIT environment for processing (12,20,22,23). As a fourth step, to prepare data for analysis we transformed the RDF files into a tabular CSV format and restructured the data into visit-centred datasets. Because outpatient encounters in SPHN-derived data lacked consistent visit identifiers, we reconstructed visit-centered records by linking timestamped clinical data elements, such as diagnoses, test results, and height and weight measurements, to outpatient encounter dates. Clinicians and hospital IT staff indicated that some clinical data elements were timestamped according to documentation or data-entry date rather than the actual encounter date. Therefore, we linked clinical data elements documented on the same day as, or after, the outpatient encounter and within 90 days. The 90-day window was chosen based on clinician and hospital IT staff input, supported by the delays observed during manual abstraction, and the distribution of diagnosis documentation delays in the received data. When several clinical data elements of the same type were available within the 90-day window, we retained the element documented closest to the outpatient encounter date. Each clinical data element was linked to only one outpatient encounter. Data transformation and reshaping workflow is detailed in **Supplementary Figure 1**. Finally, we iteratively evaluated the transformed datasets for completeness, coding consistency, and structural validity, and requested revised extracts until we obtained analysis-ready datasets. We recorded the time required to obtain analysis-ready SPHN-derived datasets and documented technical, organizational, and governance-related challenges encountered during data acquisition.

### Data analysis

#### Quantity of clinical data

We compared the number and types of respiratory-related clinical events occurring after the SPAC baseline visit between SPHN-derived data and manually abstracted or self-reported SPAC data. Events included pneumology outpatient visits, respiratory-related ED visits, and hospitalizations. Respiratory-related emergency department (ED) visits were identified from available free-text diagnostic information using a predefined keyword-matching algorithm.

The ED keyword list was developed to capture visits corresponding to the SPAC follow-up question on visits due to cough, wheezing, breathing problems, or asthma, and was refined with hospital clinicians, hospital IT staff, and SPAC study team members during iterative data-transfer quality checks. Respiratory-related hospitalizations were identified separately using ICD-10 codes for the primary diagnosis, including J00–J99, R05, and R06. Detailed information on the development of the keyword lists, the matching criteria, and the manual assessment of potential misclassification is provided in the **Supplementary Methods**. We obtained the frequencies of self-reported respiratory-related ED visits and hospitalizations from annual SPAC follow-up questionnaires. Parents were asked how often, in the preceding 12 months, their child had received care because of cough, wheezing, or asthma, including **(i)** emergency visits to a hospital emergency department without overnight stay and **(ii)** unplanned hospitalizations with one or more overnight stays. To enable quantitative comparison with SPHN-derived event counts, we converted self-reported frequencies (“none”, “once”, “twice”, “2–3”) into numeric midpoint values (0, 1, 2, and 2.5). We calculated event rates standardized per 100 person-years of follow-up time. Follow-up time was defined separately for each dataset: for SPHN-derived data, from the SPAC baseline visit to 31 December 2023 (the end of the SPHN-derived data period); for self-reported data, as the total time interval between returned follow-up questionnaires.

#### Completeness of clinical information

We compared the availability of diagnoses, anthropometric measurements, allergy testing results, and lung-function test values between manually abstracted and SPHN-derived data at SPAC baseline and pneumology follow-up visits. Additionally, we described the proportion of available clinical variables by center for respiratory-related ED visits and hospitalizations captured by SPHN-derived data.

#### Agreement and concordance between data sources

We compared the number of respiratory-related ED visits and hospitalizations recorded in SPHN-derived data with those self-reported for each 12-month follow-up period. Among participants with available self-reported data, we quantified agreement using positive, negative, and absolute percentage agreement and assessed concordance using quadratic weighted Cohen’s kappa, which accounts for chance agreement and the degree of disagreement.

We assessed agreement in selected variables between SPHN-derived and manually abstracted SPAC clinical data at the SPAC baseline visit. Specifically, we evaluated agreement for visit dates (exact matches and median absolute date differences), similarity in the diagnosis recorded in free text using the Jaccard similarity index, and sex using Cohen’s kappa. For free-text diagnoses, we defined exact matches as diagnosis pairs that were identical after basic text standardization (Jaccard similarity of 1). For non-exact pairs, we used hospital-specific Jaccard similarity thresholds, selected after manually reviewing diagnosis pairs to identify the lowest score that still represented the same clinical information. We used hospital-specific thresholds because free-text diagnoses differed between hospitals in structure, length, wording, and administrative detail. The final thresholds were 0.30 for Hospital A and 0.40 for Hospital B; we considered pairs below these thresholds discordant. We assessed agreement of continuous clinical data, such as height (cm), weight (kg), forced expiratory volume in one second (FEV₁, L), and forced vital capacity (FVC, L), using concordance correlation coefficients (CCC) and Deming regression.

We conducted all analyses in R version 4.1.1 using RStudio within BioMedIT, applying the CRAN packages *DescTools* and *MethComp* (24,25).

## RESULTS

Obtaining analysis-ready SPHN-derived datasets required approximately 24 months of work and extensive coordination across institutions, reflecting both institutional approval processes and technical challenges in transforming routinely collected clinical data into research-ready datasets. Compared with manually abstracted SPAC study data, which included clinical information from 1,971 participants across three centres (Hospital A: 517; Hospital B: 558; Hospital C: 896), SPHN-derived data were available for a smaller subset of participants. Analysis-ready SPHN-derived data were ultimately obtained for 1,075 participants from two centres (Hospital A: 517; Hospital B: 558); data from the third center (Hospital C) could not be transformed into an analysis-ready format within the 24-month study period. Achieving analysis-ready SPHN-derived datasets required completion of institutional approvals and data-sharing agreements, followed by coordination with hospital IT teams, iterative data validation, and extensive data processing by postdoctoral researchers and a PhD student **(Box 1).** Despite this effort, several clinically relevant variables could not be automatically extracted through the SPHN. Availability of extractable variables differed by center and variable type, reflecting differences in local clinical information systems that are primarily designed for clinical documentation rather than for research. Several clinically relevant variables could not be consistently extracted across centres, including outpatient treatment information and skin prick test results. In addition, some variables routinely collected in SPAC, such as family background and reasons for referral, were not available through existing SPHN Concepts **(Table 1).**

**Table 1.** Overview of information available in the Swiss Paediatric Airway Cohort (SPAC) study: manually abstracted clinical data, self-reported data from questionnaires, and automatically extracted routine hospital data acquired through the Swiss Personalized Health Network (SPHN-derived data).

| Type of information | SPAC study data |  | SPHN-derived data |
| --- | --- | --- | --- |
|  | Manually abstracted data<br>(from medical records and lung<br>function /laboratory test reports) | Self-reported data<br>(baseline and annual follow-up<br>questionnaires) | Automatically extracted from EHR and<br>lung function test devices |
| **Personal data and medical history** |  |  |  |
| Socio demographic | Sex; date of birth | Sex; date of birth; parental<br>education; address; migration<br>background | Sex; nationality (SNOMED CT); date of<br>birth; Swiss-SEP index<sup>1</sup> |
| Personal and family medical<br>history | — | Personal and family history of<br>respiratory and allergic diseases | — |
| Symptoms and environmental<br>exposures | Free-text notes on respiratory<br>symptoms, exercise-related symptoms,<br>smoking, allergen exposure | Structured responses on<br>respiratory and exercise-related<br>symptoms; smoking; allergen<br>exposure | — |
| **Clinical information** |  |  |  |
| Type of clinical event | SPAC baseline visits and at least one<br>pneumology follow up visit collected | Cumulative respiratory-related ED<br>visits and hospitalizations in<br>previous 12 months | All pneumology outpatient visits,<br>respiratory-related ED visits, and<br>respiratory-related hospitalizations<br>(SNOMED CT) |
| Reason for referral | Free text | — | — |
| Date of clinical visit | In DD.MM.YYYY format | — | In DD.MM.YYYY format |
| Diagnosis | Free-text documentation from medical<br>records and categorical diagnostic<br>terms (e.g., asthma, rhinitis, infection) | Previously reported diagnoses and<br>comorbidities | Free-text diagnoses for outpatient and<br>ED visits; ICD-10 codes (J00–J99; R05–<br>R06) for hospitalizations |
| Anthropometric measurements | Height and weight measurements<br>from medical records and lung-function test reports | — | Height and weight from structured EHR<br>entries (where available) and from lung-function test devices |
| Treatment information | Drug prescription and non-drug<br>therapies (i.e., allergen avoidance) | Current and previous medications;<br>non-pharmacological therapies | Available only for hospitalization events |
| Allergy tests | Skin prick test; total/specific IgE | History of allergy testing | Total and specific IgE results (LOINC) <sup>2</sup> |
| Lung function test | Spirometry, whole-body plethysmography, bronchoprovocation test, BDR, FeNO, nasal FeNO, MBW | History of lung function tests | Spirometry, whole-body plethysmography, BDR, bronchoprovocation test, FeNO <sup>2</sup> |
| Additional tests | X-rays, bronchoscopy, microbiology, blood cell counts, sleep studies | — | — |

<sup>1</sup> Swiss neighborhood index of socioeconomic position (Swiss-SEP): Panczak R, et al. *Health & Place*. 2012;18(4):734–741.
<sup>2</sup> FeNO and specific IgE results available only for Hospital A.
**Abbreviations:** BDR = bronchodilator response; FeNO = fractional exhaled nitric oxide; MBW = multiple breath washout; ED = emergency department; IgE = immunoglobulin E; ICD-10 = International Classification of Diseases, 10th revision; LOINC = Logical Observation Identifiers Names and Codes; SNOMED-CT = Systematized Nomenclature of Medicine – Clinical Terms.

### Box.1 Challenges encountered during acquisition and preparation of SPHN-derived data in the SPHN-SPAC demonstrator project.

Several challenges limited the availability and usability of SPHN-derived data in the SPHN–SPAC project:

#### Heterogeneous hospital systems and documentation

EHR structures, coding systems, and documentation practices varied within and between hospitals, with clinically relevant information distributed across multiple subsystems.

#### Limited extractability of key variables

Only a subset of variables collected manually in SPAC could be reliably extracted using current SPHN concepts. Particularly for outpatient care key clinical variables were missing (e.g. treatment information, outpatient diagnoses, skin prick test results).

#### RDF format and limited contextual metadata

SPHN-derived data were delivered in RDF format, requiring substantial transformation to generate analyzable, visit-centered datasets. Limited contextual metadata hindered linkage of clinical information to specific outpatient encounters.

#### Centre-specific assumptions and transformations

Reconstruction of outpatient visits relied on center-specific assumptions, using timestamps that reflected data entry rather than clinical encounters.

#### Extended implementation timeline

Data acquisition, transformation, and validation required approximately 24 months of sustained effort, including repeated revisions of data extracts and iterative validation with clinicians, hospital IT teams, and SPHN data engineers.

#### Comparison with manual data abstraction

Manual SPAC data collection involved recurring on-site visits and abstraction of baseline and first follow-up visits, with an estimated research team cost of approximately CHF 97,000. Preparation of SPHN-derived data required a larger upfront investment (approximately CHF 242,000, excluding hospital IT and governance costs). Achieving comparable longitudinal coverage through manual abstraction would have required an estimated CHF 167,000 and approximately eight additional months of work. In addition, manual abstraction introduces potential for transcription errors as workload increases, reflecting limitations inherent to manual data handling.

**Abbreviations:** RDF= Resource Description Framework.

Preparing SPHN-derived data for analysis required transformation from RDF format into tabular datasets organized by clinical visits. Outpatient data lacked unique visit identifiers; therefore, we assigned anthropometric and lung function measurements, diagnoses and laboratory results to distinct clinical visits based on timestamps as described in the methods.

### Data quantity

The SPHN-derived dataset included more clinical visits (baseline plus follow-up visits) than the manually abstracted SPAC data at both centres (Hospital A: 1,963 vs 1,049; Hospital B: 2,343 vs 1,010; **Figure 2**). This was because manual abstraction of clinical data in SPAC was limited to the enrolment visit and one subsequent pneumology follow-up visit per child for feasibility, whereas SPHN-derived data included all subsequent pneumology outpatient visits and lung function tests recorded during routine care over the entire observation period. Correspondingly, the proportion of children with at least one follow-up visit increased from 61% to 77% at Hospital A and from 48% to 81% at Hospital B. Over the study period, SPHN-derived data recorded more children with recorded respiratory-related ED visits than self-reported data (Hospital A: 53 vs 44; Hospital B: 83 vs 58). However, the rate of ED visits was higher in the self-reported data (Hospital A: 8.7 vs 3.8 per 100 person-years; Hospital B: 10 vs 5.3). For hospitalizations, the number of children with recorded respiratory-related hospitalizations was comparable between datasets (Hospital A: 15 vs 18; Hospital B: 17 vs 20), and hospitalization rates were higher in the self-reported data (Hospital A: 3 vs 1; Hospital B: 2 vs 1 per 100 person-years).

**Figure 2.**
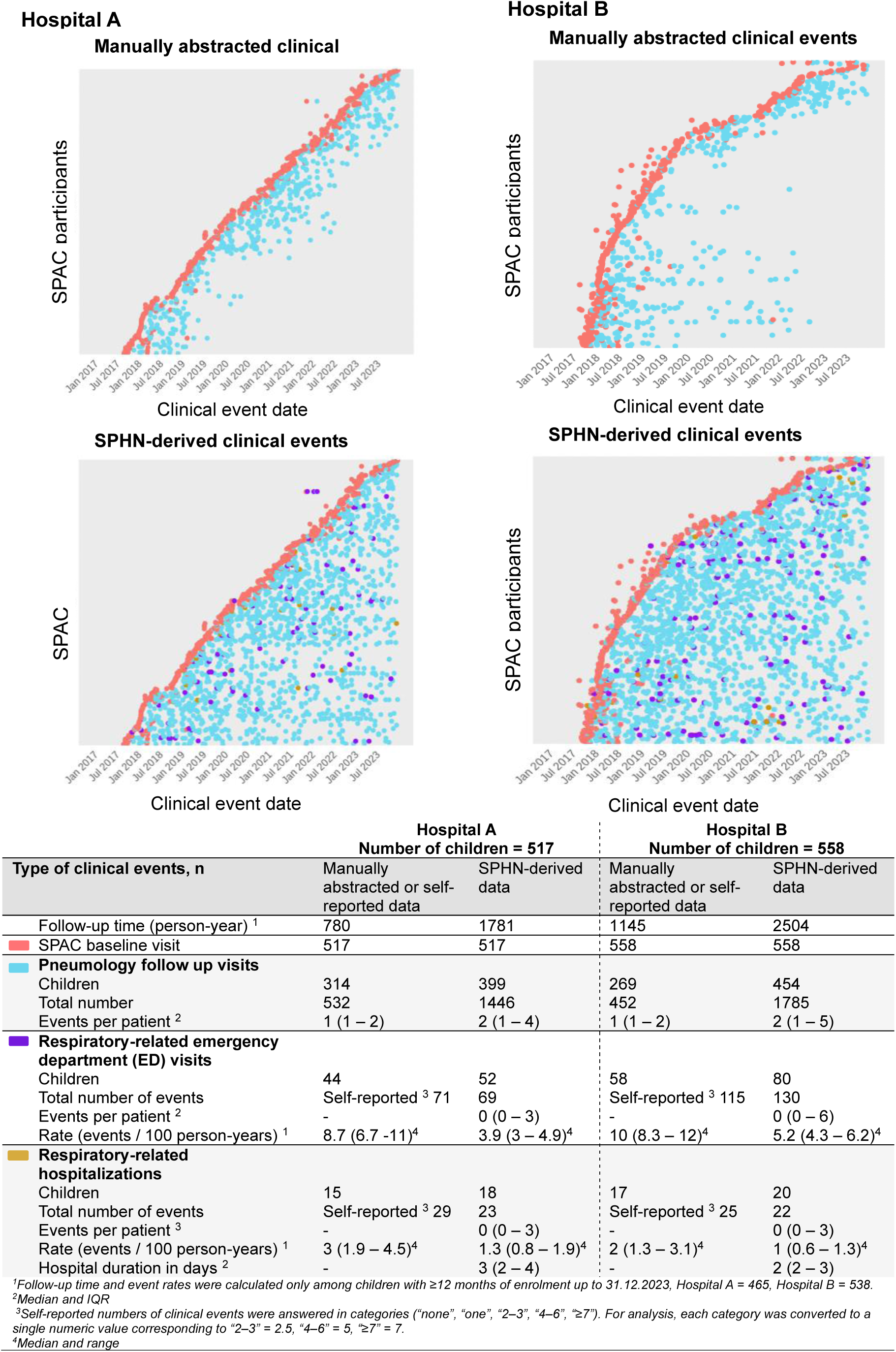
Respiratory-related clinical event information derived from manually abstracted medical records or self-reported questionnaire data and SPHN-derived data, for each hospital.

### Data completeness

Completeness, defined as the availability of clinical information for each type of clinical encounter (pneumology outpatient visit, ED visit, or hospitalization), varied by hospital and encounter type (**Table 2**). At the baseline visit, manually abstracted SPAC data showed near-complete availability of anthropometric measurements obtained from clinical letters (height: 98–99%; weight: 96–99%). Similarly, SPHN-derived data showed high completeness for the same variables (>95%) at Hospital B, whereas completeness at Hospital A was substantially lower (62–63%). For pneumology follow-up visits, spirometry measurements were available for a substantial proportion of encounters in SPHN-derived data (Hospital A: 75%; Hospital B: 97%), representing approximately twice the number of spirometry measurements available through manually abstracted data for the same participants. However, at the baseline visit, more spirometry measurements were available in manually abstracted data than in SPHN-derived data for Hospital A.

**Table 2.** Completeness of clinical information from manually abstracted and SPHN-derived clinical data at SPAC baseline visit and pneumology follow-up visits.

| Availability of clinical information, n (%) |  |  |  |  |
| --- | --- | --- | --- | --- |
| Clinical information | Hospital A n = 517 |  | Hospital B n = 558 |  |
|  | Manually abstracted data | SPHN-derived data | Manually abstracted data | SPHN-derived data |
| **SPAC baseline visit** |  |  |  |  |
| Height<sup>1</sup> | 508 (98) | 322 (62) | 533 (96) | 545 (98) |
| Weight<sup>1</sup> | 511 (99) | 325 (63) | 535 (96) | 554 (99) |
| Diagnosis | 513 (99) | 509 (99) | 556 (99) | 556 (99) |
| Serum IgE | 20 (4) | 73 (14) | 249 (44) | 358 (64) |
| Spirometry | 421 (82) | 354 (68) | 456 (82) | 455 (82) |
| BDR test | 170 (32) | 67 (19) | 327 (72) | 329 (59) |
| Body plethysmography | 366 (72) | 316 (61) | 245 (44) | 251 (45) |
| FeNO | 395 (77) | 345 (67) | 506 (91) | Not available |
| Height from lung function device<sup>2</sup> | 408 (79) | 390 (75) | 455 (82) | 455 (82) |
| Weight from lung function device<sup>2</sup> | 408 (79) | 390 (75) | 455 (82) | 455 (82) |
| **Pneumology follow up visits** |  |  |  |  |
| Number of clinical events extracted | 533 | 1446 | 452 | 1785 |
| Number of patients | 314 (60) | 398 (71) | 269 (48) | 454 (81) |
| Events per patient, median (IQR) | 1 (1–2) | 2 (1–4) | 1 (1–2) | 2 (1–5) |
| Diagnosis | 447 (84) | 1389 (96) | 358 (79) | 1749 (98) |
| Height<sup>1</sup> | 433 (81) | 1092 (75) | 339 (75) | 1714 (96) |
| Weight<sup>1</sup> | 434 (81) | 1121 (77) | 342 (76) | 1748 (98) |
| Spirometry | 387 (89) | 1084 (75) | 285 (73) | 1402 (97) |
| BDR test | 128 (35) | 267 (25) | 143 (63) | 1039 (58) |
| Body plethysmography | 356 (83) | 1045 (72) | 93 (25) | 692 (48) |
| Challenge test | 49 (12) | 107 (7) | 26 (7.3) | 47 (3.2) |
| FeNO | 357 (85) | 1103 (75) | 251 (56) | Not available |
| Height from lung function device<sup>2</sup> | 359 (67) | 1223 (84) | 218 (48) | 1406 (97) |
| Weight from lung function device<sup>2</sup> | 359 (67) | 1223 (84) | 218 (48) | 1406 (97) |

<sup>1</sup> Height and weight extracted from structured anthropometric fields in the EHR.
<sup>2</sup> Height and weight extracted from lung function device.
**Abbreviations:** BDR = bronchodilator response test; FeNO = fractional exhaled nitric oxide; Challenge test = Methacholine bronchoprovocation test

FeNO measurements were available for 67% of SPAC baseline visits at Hospital A and 75% of follow-up visits but were not available for extraction at Hospital B at the time of data analysis due to ongoing mapping of lung function data to SPHN standards. Treatment information from SPHN-derived data could not be extracted for outpatient pneumology visits but was partially available for ED visits and hospitalizations, with substantial variation between hospitals (Hospital A: 21% and 78%; Hospital B: 98% and 91%, respectively; Supplementary Table 2).

### Agreement and concordance between datasets

In 12-month follow-up periods with available self-reported data, absolute and negative agreement between SPHN-derived and self-reported data for respiratory-related ED visits and hospitalizations were very high, whereas positive agreement and concordance measured using weighted kappa statistics were moderate (**Table 3**). At Hospital A, percentage agreement was 94% for respiratory-related ED visits and 98% for hospitalizations, with corresponding weighted κ values of 0.43 and 0.55. At Hospital B, percentage agreement was 90% and 98%, with κ values of 0.23 for ED visits and 0.21 for hospitalizations. In follow-up periods without self-reported questionnaire data, most periods had no ED visits or hospitalizations recorded in SPHN-derived data (Hospital A: 96% without ED visits and >99% without hospitalizations; Hospital B: 98% without ED visits and >99% without hospitalizations). SPHN-derived data nevertheless identified a small number of 12-month periods with respiratory-related ED visits (Hospital A: 32 of 777 periods; Hospital B: 52 of 1,095 periods) and hospitalizations (Hospital A: 7 of 789 periods; Hospital B: 6 of 1,100 periods) among participants for whom self-reported data were unavailable.

**Table 3.** Agreement and concordance between self-reported healthcare utilization from questionnaire data and SPHN-derived data on respiratory-related emergency department (ED) visits and hospitalizations, across 12-month follow-up periods.

**Hospital A- Respiratory-related ED visits**
|  |  | SPHN-derived data |  |  |  |
| --- | --- | --- | --- | --- | --- |
| Self-reported |  | 0 | 1 | ≥ 2 | Total |
|  | 0 | 703 | 6 | 1 | 710 |
|  | 1 | 26 | 14 | 0 | 40 |
|  | ≥ 2 | 6 | 2 | 2 | 10 |
|  | NA | 745 | 28 | 4 | 777 |
|  | Total | 1480 | 50 | 7 | 1537 |
| Percent agreement = 94 %<br>Positive agreement = 47 %<br>Negative agreement = 97 %<br>Weighted kappa (complete cases) = 0.43 (0.28-0.56) |  |  |  |  |  |

**Hospital B- Respiratory-related ED visits**
|  |  | SPHN-derived data |  |  |  |
| --- | --- | --- | --- | --- | --- |
| Self-reported |  | 0 | 1 | ≥ 2 | Total |
|  | 0 | 1024 | 24 | 7 | 1055 |
|  | 1 | 46 | 8 | 1 | 55 |
|  | ≥ 2 | 14 | 6 | 3 | 23 |
|  | NA | 1043 | 46 | 6 | 1095 |
|  | Total | 2127 | 84 | 17 | 2228 |
| Percent agreement = 91%<br>Positive agreement = 28%<br>Negative agreement = 96%<br>Weighted kappa (complete cases) = 0.23 (0.13-0.33) |  |  |  |  |  |

**Hospital A- Respiratory-related hospitalizations**
|  |  | SPHN-derived data |  |  |  |
| --- | --- | --- | --- | --- | --- |
| Self-reported |  | 0 | 1 | ≥ 2 | Total |
|  | 0 | 727 | 4 | 0 | 731 |
|  | 1 | 6 | 6 | 0 | 12 |
|  | ≥ 2 | 2 | 2 | 1 | 5 |
|  | NA | 7182 | 6 | 1 | 789 |
|  | Total | 1517 | 18 | 2 | 1537 |
| Percent agreement = 98 %<br>Positive agreement = 60 %<br>Negative agreement = 99 %<br>Weighted kappa (complete cases) = 0.55 (0.30-0.74) |  |  |  |  |  |

**Hospital B- Respiratory-related hospitalizations**
|  |  | SPHN-derived data |  |  |  |
| --- | --- | --- | --- | --- | --- |
| Self-reported |  | 0 | 1 | ≥ 2 | Total |
|  | 0 | 1100 | 7 | 1 | 1108 |
|  | 1 | 14 | 3 | 0 | 17 |
|  | ≥ 2 | 2 | 1 | 0 | 3 |
|  | NA | 1094 | 6 | 0 | 1100 |
|  | Total | 2201 | 17 | 1 | 2228 |
| Percent agreement = 98 %<br>Positive agreement = 25%<br>Negative agreement = 99%<br>Weighted kappa (complete cases) = 0.21 (0.04-0.39) |  |  |  |  |  |

- Cells represent the number of 12-month follow-up periods with corresponding self-reported and SPHN-derived hospital data clinical events. Among children with at least 12 months of follow up (Hospital A = 465 ; Hospital B = 538) - For self-reported data = One returned follow-up questionnaire equals 1 follow-up period - For SPHN-derived data = clinical events were assigned to the nearest 12-month period. - -NA = Missing self-reported data (no returned questionnaire). - Percent agreements and linearly weighted Cohen's $\kappa$ (95% CI) calculated for available time periods with available data in both datasets: - Hospital A: ED (n = 765), hospitalizations (n = 753), and Hospital B: ED (n = 1,134), hospitalizations (n = 1,129)

Concordance between SPHN-derived data and manually abstracted SPAC clinical data at the SPAC baseline visit was high across all evaluated variables **(Table 4).** Visit dates matched exactly for 95% of the baseline visits at Hospital A and 99% at Hospital B, with a median difference of 0 days. Sex showed near-perfect concordance (κ = 0.95–0.96). After text standardization, we observed exact diagnosis matches in 308/517 baseline visits (60%) at Hospital A and 323/558 visits (58%) at Hospital B. Including non-exact pairs classified as clinically similar using hospital-specific Jaccard thresholds, diagnoses were concordant in 505/517 visits (98%) and 552/558 visits (99%), respectively (Table 4 and Supplementary Table 6). Diagnoses recorded as free text showed a high degree of similarity between datasets, with similar diagnostic wording identified in more than 95% of visits at both centres (Hospital A: 504/517; Hospital B: 552/558). Agreement for anthropometric measurements and spirometry outcomes was also very high (CCC height: 0.97–0.99; CCC weight: 0.91–0.99; CCC FEV₁: 0.99–0.99; CCC FVC: 0.99–0.99). Visual assessments of concordance are shown in **Supplementary Figures 2–8.**

**Table 4.**
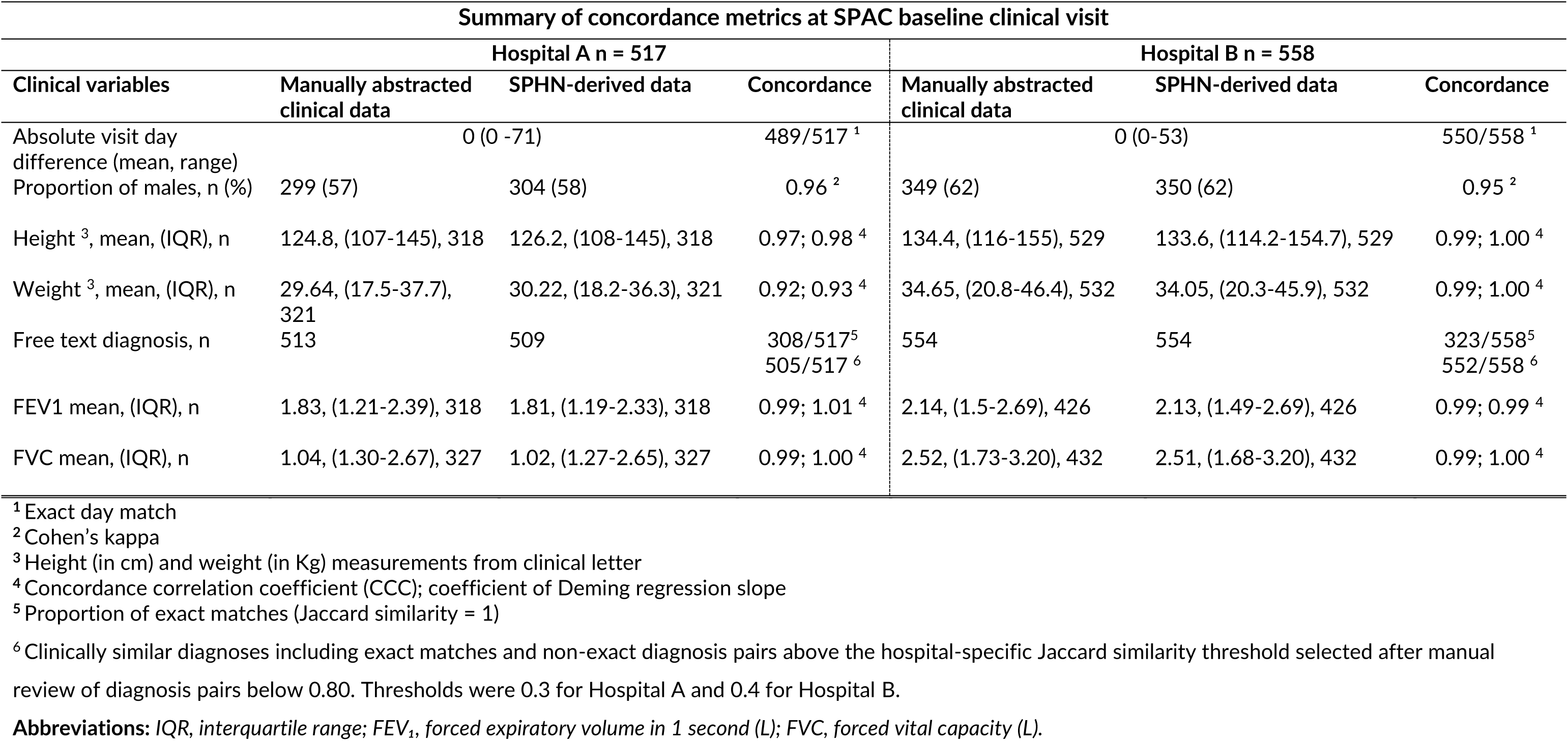
Summary of concordance metrics for key clinical variables between manually abstracted medical records and SPHN-derived clinical.

## DISCUSSION

In this feasibility study, SPHN-derived routine clinical data substantially increased the availability of longitudinal clinical information compared with manual data abstraction in the Swiss Paediatric Airway Cohort (SPAC), while maintaining very high agreement for key structured clinical variables. Obtaining analysis-ready SPHN-derived datasets required substantial time, human resources, and coordination, taking approximately 24 months and multiple rounds of extraction, validation, and restructuring across institutions. Once obtained, SPHN-derived data substantially increased the availability of respiratory-related clinical information, capturing nearly twice as many pneumology outpatient visits (Hospital A: 1,963 vs 1,049; Hospital B: 2,343 vs 1,010) and more spirometry measurements than manually abstracted SPAC data (Hospital A: 1,438 vs 808; Hospital B: 1,857 vs 741), reflecting the inclusion of all routinely documented encounters during the observation period. Agreement between SPHN-derived and manually abstracted SPAC clinical data was very high for key structured variables, with exact agreement for most visit dates and near-perfect concordance for spirometry measurements (CCC for FEV₁ and FVC ≈0.99), indicating that structured clinical data recorded in hospital systems can be reliably extracted through the SPHN framework.

Despite these advantages, obtaining analysis-ready SPHN-derived datasets required considerable upfront effort and revealed important limitations for broader research use. Manual data collection in SPAC involved recurring on-site visits and participant-level abstraction of baseline and first follow-up visits, requiring several hundred hours of work that scaled with cohort size. In contrast, although SPHN-derived data ultimately provided substantially more data points, the initial effort required to obtain, harmonize, and prepare these data for analysis exceeded that required for manual abstraction during the same study period. In addition, successful automated extraction was largely limited to structured clinical variables, with data availability and completeness varying across hospitals, reflecting differences in local documentation practices and information systems. For instance, important information like diagnosis of treatment were not available in a structured (analyzable) way for many outpatient visits.

Similar to large EHR-based research networks such as PEDSnet and PCORnet, the use of routinely collected clinical data in our study required substantial upfront investment in data harmonization and validation (17,26–29). In our study, the challenges we observed reflected both the structural requirements of the SPHN framework and differences in local clinical information systems across centres. Although clinically relevant variables were present in hospital records, they could not always be successfully extracted through the SPHN framework. Automated extraction pipelines require data to be stored and labelled according to specific terminology standards and documentation structures; when local documentation deviated from these specifications, variables were not extracted, only partially available, or incorrectly mapped. For example, FeNO measurements were extracted successfully at Hospital A, whereas at Hospital B the same pipeline returned nasal FeNO rather than exhaled FeNO because of local coding and mapping differences.

We also found that clinical data were documented and stored differently across hospitals, resulting in variable availability and completeness of automatically extracted data and requiring substantial processing before analysis. These findings suggest that barriers to broader use of SPHN-derived data arise from both the lack of uniformly available structured data and heterogeneity in clinical documentation practices across centres, as well as from the substantial effort required to transform extracted data into analysis-ready datasets.

Differences between SPHN-derived and self-reported emergency department (ED) visits and hospitalizations likely reflect differences in how clinical events are captured by the two datasets. SPHN-derived data are restricted to encounters recorded within participating hospitals and linked to specific dates and therefore did not capture ED visits or hospitalizations at non-participating hospitals. In contrast, self-reported data could include care received at other healthcare facilities. While hospitalizations usually occurred at the same hospital where participants received outpatient care, because other hospitals were located farther away, emergency visits could also have occurred at ambulatory walk-in clinics or paediatric practices. Self-reported events may also have been affected by recall error and differences between the questionnaire completion date, questionnaire return date, and the SPHN-derived 12-month follow-up periods, resulting in some events being assigned to different follow-up periods.

High absolute and negative agreement should therefore be interpreted cautiously because most follow-up periods contained no respiratory-related ED visits or hospitalizations in either dataset. Positive agreement and κ statistics were lower, indicating that when at least one dataset recorded an event, the two datasets did not always classify the same follow-up period as event-positive. This discrepancy was more pronounced for ED visits than for hospitalizations, likely because ED visits more often occurred outside the participating hospitals and being less severe, may have been recalled less accurately by parents (30).

The comparable total numbers of hospitalizations identified by self-report and SPHN-derived data, 29 versus 23 in Hospital A and 25 versus 22 in Hospital B, suggest that the lower κ values for hospitalizations may partly reflect differences in event timing rather than differences in overall event capture. SPHN-derived data also captured respiratory-related ED visits and hospitalizations among children with missing self-reported data due to unreturned questionnaires, highlighting the value of routinely extracted clinical data to complement participant-reported information in longitudinal cohort studies.

### Strengths and limitations

This study has several limitations. Preparing SPHN-derived data required substantial time and coordination, and only two of the three participating hospitals delivered analysis-ready datasets within the 24-month study period. This appeared to reflect differences in whether local EHR data could be extracted, mapped to SPHN concepts, and transformed into an analyzable format within the study period, rather than absence of relevant clinical information. The two centres that delivered data were university hospitals with dedicated IT and clinical support, whereas the third was a cantonal hospital with fewer dedicated resources and was participating in this type of extraction for the first time. Although we did not formally compare institutional systems, these differences in prior experience and research support may have contributed to differences in timely data delivery. Creating visit-centered datasets required extensive transformation and harmonization, which may have introduced some misclassification or data-processing artefacts. The fixed 90-day linkage window allowed us to link delayed documentation such as diagnoses entered into the EHR after the clinical encounter to the relevant visit but may have introduced ambiguity for children with closely spaced visits. To minimize this risk, we linked only the clinical data element closest to the outpatient encounter. Nevertheless, some residual linkage error may remain, particularly for children with frequent visits. Additionally, keyword-based definitions for emergency department visits may have led to false-positive or false-negative classifications when diagnostic text was ambiguous, incomplete, or not extractable from the SPHN-derived data. Nevertheless, the manual reviews indicated that residual misclassification was likely negligible. In addition, our evaluation was limited to clinical variables represented by the SPHN concepts selected for extraction and did not assess the feasibility of using unstructured clinical text. Finally, because our study included only a paediatric respiratory cohort from two tertiary centres with well-developed informatics infrastructure, the feasibility of using SPHN-derived data may be overestimated compared with other Swiss hospitals. Despite these limitations, our study has several strengths. To our knowledge, it provides the first detailed evaluation of the feasibility and performance of SPHN-derived paediatric clinical data in a real-world multicenter research setting. Including hospitals with different EHR systems and documentation practices allowed evaluation under realistic interoperability conditions. By comparing automatically extracted, manually abstracted, and self-reported data for the same participants, we assessed validity, completeness, and the complementary value of the different datasets. The study also benefited from the national SPHN framework, which provides shared semantic models and governance structures, and from the secure BioMedIT infrastructure, which enabled secure and reproducible data handling.

### Implications and future directions

The challenges identified in this demonstrator study illustrate why the broader and scalable use of SPHN-derived routine clinical data in paediatric research is not yet straightforward **(Box 2).** At the same time, the extraction pipelines and data-processing workflows developed in this project provide a reusable foundation that may reduce the time and resources required for future implementations and facilitate similar applications in other cohort studies and clinical domains using the SPHN infrastructure. The challenges observed in this study largely reflect the practical consequences of current clinical documentation practices and the structural requirements of the SPHN framework. In particular, variability in outpatient documentation and the absence of explicit visit-level identifiers require hospital-and project-specific assumptions and workflows, limiting direct reuse across centres and studies. In addition, a substantial proportion of clinically relevant information remains recorded in unstructured or semi-structured free-text notes, which cannot be fully captured by current SPHN-derived data and therefore limits data completeness. Extending the SPHN framework to support the extraction and standardization of information from free-text clinical documentation, for example through natural language processing approaches, could substantially improve data completeness and reduce reliance on manual data collection (31,32). Applying SPHN-derived data to additional centres or clinical populations will therefore require careful evaluation of local data extractability and documentation practices. This requires dedicated IT support and close collaboration with clinicians and researchers to identify mapping issues and perform iterative quality checks. Centres undergoing SPHN-based data harmonization and extraction for the first time may require additional start-up support, particularly when local experience with SPHN concepts and RDF-based data structures is limited. At the SPHN level, practical examples, code, and guidance showing how hospitals using similar EHR systems have extracted and mapped their data to SPHN concepts could make implementation easier for new centres. Researchers would also benefit from examples and scripts showing how RDF-based SPHN data can be transformed into tabular datasets for analysis in common statistical software. Ultimately, broader implementation of SPHN-derived data will depend on alignment between research questions, hospital data infrastructure, and clinical documentation practices, rather than on technical data extraction alone.

## Conclusions

SPHN-derived routine clinical data can complement manual data collection in paediatric cohort studies by increasing the availability of clinical information and improving follow-up completeness while maintaining high validity for structured variables. However, they cannot yet replace manual data collection, as broader use remains constrained by heterogeneous clinical documentation across hospitals, challenges in linking outpatient data to specific visits, and the absence of analysis-ready data delivery formats. Addressing these limitations through improved harmonization of clinical documentation practices and development of standardized analysis-ready SPHN datasets will be essential for broader use. Building on the pipelines and workflows developed in this demonstrator project, and with further improvements in documentation harmonization and data delivery formats, SPHN-derived data have the potential to substantially reduce manual workload, improve research efficiency, and strengthen the completeness and robustness of evidence generated in paediatric observational research.

### Box 2. Conditions for broader reuse of SPHN-derived routine clinical data in paediatric research

To enable broader, scalable use of SPHN-derived routine clinical data, several challenges identified in this study need to be addressed at the hospital, SPHN infrastructure, and research levels.
**Hospital level:**

- Standardized clinical documentation.

∎ The reliable reuse of SPHN-derived data depends on consistent documentation of diagnoses, treatments, and test results within electronic health records in participating clinics. In this study, heterogeneity in how and where information was recorded within and between hospitals substantially limited the extractability of clinically relevant variables, despite the use of standardized SPHN concepts. Greater harmonization of clinical documentation, particularly for outpatient care, is therefore a prerequisite for scalable reuse of routinely collected data.
- Dedicated IT capacity

∎Reusing SPHN-derived data requires local expertise to extract routine clinical data, identify where relevant information is stored within local EHR systems, and map these data to SPHN concepts. Dedicated hospital IT staff should work closely with clinicians and researchers to support preliminary data transfers, repeated quality checks, and early detection of extraction or mapping problems. Centres doing this for the first time may benefit from shared code or mapping instructions from hospitals using similar EHR systems

**SPHN level:**

- Centralized transformation and research-ready outputs

∎SPHN data are delivered in Resource Description Framework (RDF) format to support semantic interoperability but require extensive, project-specific transformation and knowledge before they can be analyzed. Centrally maintained transformation workflows, visit-identification logic, or optional analysis-ready visit-centered outputs would reduce technical burden and improve reproducibility across studies.
- Explicit visit-level identifiers for outpatient care

∎Current SPHN concepts lack consistent visit-level identifiers for outpatient clinical events. As a result, outpatient visits must be reconstructed by linking time-stamped clinical events, with timestamps often reflecting data entry rather than the actual date of care and requiring center-specific assumptions. Introducing explicit visit-level identifiers for outpatient encounters would simplify data processing, reduce ambiguity, and improve comparability across centres and studies.
- Improved capture of clinically relevant information in free text

∎A substantial proportion of routine paediatric care is documented only in unstructured or semi-structured free-text notes. Extending the SPHN framework to support extraction and standardization of free-text information, for example through natural language processing, would improve data completeness and reduce reliance on manual abstraction.

**Researcher level:**

- Early integration of data constraints into study design

∎Even with improvements at the hospital and SPHN levels, effective use of SPHN-derived data requires researchers to explicitly account for the structural characteristics and limitations of routine clinical data at the study design stage. Early collaboration with clinicians, hospital IT teams, and data providers is essential to identify which variables are reliably extractable, how clinical events are represented, and which research questions can be addressed using SPHN-derived data.

## DECLARATIONS

## Data Availability

All data produced in the present study are available upon reasonable request to the authors

## Acknowledgements

We thank the hospital IT teams and data management staff at Inselspital, Bern University Hospital (Bern), the University Children’s Hospital Zurich (Zurich), and the Lucerne Cantonal Hospital (Lucerne) for their technical support, including Beat Bangerter, Simeon Haefliger, and Mathias Grassner, whose collaboration enabled the acquisition of SPHN-derived data. We also acknowledge the clinicians, hospital staff, and study teams involved in the SPHN–SPAC project and PedNet Bern for supporting data collection in Bern, as well as the members of the SPAC study team. Finally, we thank the families who took part in the SPAC study. This paper incorporates suggestions from AI tools, including Grammarly and ChatGPT (OpenAI), to improve grammar and readability. All scientific content and conclusions are the responsibility of the authors.

## Ethical considerations

The Cantonal Ethics Committee of Bern (KEB 2016-02176) granted ethical approval for the SPAC study and SPHN-SPAC demonstrator project. Written informed consent was obtained from parents or legal guardians for all SPAC participants, and directly from participants aged ≥14 years. All data handling complied with Swiss data protection regulations and was conducted within secure infrastructures (REDCap and BioMedIT).

## REFERENCES

1. Ahmed MF, Galfo A, Huggins W, Paddock LE, Stroup AM, Malhotra J. Challenges of Medical Record Abstraction in a Long-Term Follow-up Study. J Regist Manag. 2021;48(3):88–91. PubMed PMID: 35413725.

2. Garza MY, Williams TB, Ounpraseuth S, Hu Z, Lee J, Snowden J, et al. Comparing Medical Record Abstraction (MRA) error rates in an observational study to pooled rates identified in the data quality literature. BMC Med Res Methodol. 2024 Dec 18;24(1):304. doi:10.1186/s12874-024-02424-x

3. Forrest CB, Margolis PA, Bailey LC, Marsolo K, Del Beccaro MA, Finkelstein JA, et al. PEDSnet: a National Pediatric Learning Health System. J Am Med Inform Assoc JAMIA. 2014 Jul;21(4):602–6. doi:10.1136/amiajnl-2014-002743 PubMed PMID: 24821737; PubMed Central PMCID: PMC4078288.

4. Walsh KE, Razzaghi H, Hartley DM, Utidjian L, Alford S, Darwar RA, et al. Testing the Use of Data Drawn from the Electronic Health Record to Compare Quality. Pediatr Qual Saf. 2021 Jul 28;6(4):e432. doi:10.1097/pq9.0000000000000432 PubMed PMID: 34345748; PubMed Central PMCID: PMC8322494.

5. Forrest CB, McTigue KM, Hernandez AF, Cohen LW, Cruz H, Haynes K, et al. PCORnet® 2020: current state, accomplishments, and future directions. J Clin Epidemiol. 2021 Jan;129:60–7. doi:10.1016/j.jclinepi.2020.09.036 PubMed PMID: 33002635; PubMed Central PMCID: PMC7521354.

6. Lewis AE, Weiskopf N, Abrams ZB, Foraker R, Lai AM, Payne PRO, et al. Electronic health record data quality assessment and tools: a systematic review. J Am Med Inform Assoc JAMIA. 2023 Sep 25;30(10):1730–40. doi:10.1093/jamia/ocad120 PubMed PMID: 37390812; PubMed Central PMCID: PMC10531113.

7. An D, Lim M, Lee S. Challenges for Data Quality in the Clinical Data Life Cycle: Systematic Review. J Med Internet Res. 2025 Apr 23;27(1):e60709. doi:10.2196/60709

8. Ozonze O, Scott PJ, Hopgood AA. Automating Electronic Health Record Data Quality Assessment. J Med Syst. 2023;47(1):23. doi:10.1007/s10916-022-01892-2 PubMed PMID: 36781551; PubMed Central PMCID: PMC9925537.

9. Lawrence AK, Selter L, Frey U. SPHN - The Swiss Personalized Health Network Initiative. Stud Health Technol Inform. 2020 Jun 16;270:1156–60. doi:10.3233/SHTI200344 PubMed PMID: 32570562.

10. Rakic M, Jaboyedoff M, Bachmann S, Berger C, Diezi M, do Canto P, et al. Clinical data for paediatric research: the Swiss approach. BMC Proc. 2021 Sep 20;15(13):19. doi:10.1186/s12919-021-00226-3

11. Armida J, Touré V, Krauss P, Unni D, Chiarugi D, Marto AB, et al. Semantic Interoperability at National Scale: The SPHN Federated Clinical Routine Dataset [Internet]. Research Square; 2025 [cited 2025 Dec 9]. Available from: https://www.researchsquare.com/article/rs-8250886/v1 doi:10.21203/rs.3.rs-8250886/v1

12. Touré V, Krauss P, Gnodtke K, Buchhorn J, Unni D, Horki P, et al. FAIRification of health-related data using semantic web technologies in the Swiss Personalized Health Network. Sci Data. 2023 Mar 10;10(1):127. doi:10.1038/s41597-023-02028-y

13. Jaboyedoff M, Rakic M, Bachmann S, Berger C, Diezi M, Fuchs O, et al. SwissPedData: Standardising hospital records for the benefit of paediatric research. Swiss Med Wkly. 2021 Dec 21;151(5152):w30069. doi:10.4414/SMW.2021.w30069

14. Mozun R, Belle FN, Agostini A, Baumgartner MR, Fellay J, Forrest CB, et al. Paediatric Personalized Research Network Switzerland (SwissPedHealth): a joint paediatric national data stream. BMJ Open. 2024 Dec 26;14(12):e091884. doi:10.1136/bmjopen-2024-091884 PubMed PMID: 39725440; PubMed Central PMCID: PMC11683899.

15. Pedersen ESL, Jong CCM de, Ardura-Garcia C, Barben J, Casaulta C, Frey U, et al. The Swiss Paediatric Airway Cohort (SPAC). ERJ Open Res. 2018 Oct 1;4(4). doi:10.1183/23120541.00050-2018

16. Harris PA, Taylor R, Thielke R, Payne J, Gonzalez N, Conde JG. Research electronic data capture (REDCap)—A metadata-driven methodology and workflow process for providing translational research informatics support. J Biomed Inform. 2009 Apr 1;42(2):377–81. doi:10.1016/j.jbi.2008.08.010

17. Bartlett VL, Dhruva SS, Shah ND, Ryan P, Ross JS. Feasibility of Using Real-World Data to Replicate Clinical Trial Evidence. JAMA Netw Open. 2019 Oct 9;2(10):e1912869. doi:10.1001/jamanetworkopen.2019.12869 PubMed PMID: 31596493; PubMed Central PMCID: PMC6802419.

18. The SPHN RDF Schema [Internet]. [cited 2026 Feb 26]. Available from: https://www.biomedit.ch/rdf/sphn-schema/sphn/2026/1

19. Coman Schmid D, Crameri K, Oesterle S, Rinn B, Sengstag T, Stockinger H, et al. SPHN - The BioMedIT Network: A Secure IT Platform for Research with Sensitive Human Data. Stud Health Technol Inform. 2020 Jun 16;270:1170–4. doi:10.3233/SHTI200348 PubMed PMID: 32570566.

20. Casarano A. SPHN Connector: Public release of our open-source tool. SPHN [Internet]. 2023 Oct 19 [cited 2025 Dec 9]. Available from: https://sphn.ch/2023/10/19/sphn-connector-public-release/

21. The SPHN RDF Schema [Internet]. [cited 2025 Dec 9]. Available from: https://www.biomedit.ch/rdf/sphn-schema/sphn/2024/2

22. Gaudet-Blavignac C, Raisaro JL, Touré V, Österle S, Crameri K, Lovis C. A National, Semantic-Driven, Three-Pillar Strategy to Enable Health Data Secondary Usage Interoperability for Research Within the Swiss Personalized Health Network: Methodological Study. JMIR Med Inform. 2021 Jun 24;9(6):e27591. doi:10.2196/27591

23. RDF 1.1 Concepts and Abstract Syntax [Internet]. [cited 2026 Feb 26]. Available from: https://www.w3.org/TR/rdf11-concepts/

24. Signorell A, Aho K, Alfons A, Anderegg N, Aragon T, Arachchige C, et al. DescTools: Tools for Descriptive Statistics [Internet]. 2025 [cited 2025 Nov 2]. Available from: https://cran.r-project.org/web/packages/DescTools/index.html

25. Carstensen B, Gurrin L, Ekstrøm CT, Figurski M. MethComp: Analysis of Agreement in Method Comparison Studies [Internet]. 2024 [cited 2025 Nov 2]. Available from: https://cran.r-project.org/web/packages/MethComp/index.html

26. Yin AL, Guo WL, Sholle ET, Rajan M, Alshak MN, Choi JJ, et al. Comparing automated vs. manual data collection for COVID-specific medications from electronic health records. Int J Med Inf. 2021 Oct 21;157:104622. doi:10.1016/j.ijmedinf.2021.104622 PubMed PMID: 34741892.

27. Ebbers T, Takes RP, Honings J, Smeele LE, Kool RB, van den Broek GB. Development and validation of automated electronic health record data reuse for a multidisciplinary quality dashboard. Digit Health. 2023 Jul 28;9:20552076231191007. doi:10.1177/20552076231191007 PubMed PMID: 37529541; PubMed Central PMCID: PMC10388626.

28. Skyttberg N, Chen R, Koch S. Man vs machine in emergency medicine – a study on the effects of manual and automatic vital sign documentation on data quality and perceived workload, using observational paired sample data and questionnaires. BMC Emerg Med. 2018 Dec 13;18:54. doi:10.1186/s12873-018-0205-2 PubMed PMID: 30545312; PubMed Central PMCID: PMC6293611.

29. Mahmoudi E, Kamdar N, Kim N, Gonzales G, Singh K, Waljee AK. Use of electronic medical records in development and validation of risk prediction models of hospital readmission: systematic review. BMJ. 2020 Apr 8;369:m958. doi:10.1136/bmj.m958 PubMed PMID: 32269037.

30. Short ME, Goetzel RZ, Pei X, Tabrizi MJ, Ozminkowski RJ, Gibson TB, et al. How accurate are self-reports? Analysis of self-reported health care utilization and absence when compared with administrative data. J Occup Environ Med. 2009 Jul;51(7):786–96. doi:10.1097/JOM.0b013e3181a86671 PubMed PMID: 19528832; PubMed Central PMCID: PMC2745402.

31. Warner JL, Levy MA, Neuss MN, Warner JL, Levy MA, Neuss MN. ReCAP: Feasibility and Accuracy of Extracting Cancer Stage Information From Narrative Electronic Health Record Data. J Oncol Pract. 2016 Feb;12(2):157–8. doi:10.1200/JOP.2015.004622

32. Cai T, Zhang L, Yang N, Kumamaru KK, Rybicki FJ, Cai T, et al. EXTraction of EMR numerical data: an efficient and generalizable tool to EXTEND clinical research. BMC Med Inform Decis Mak. 2019 Nov 15;19(1):226. doi:10.1186/s12911-019-0970-1

