## Supplementary material for "Feasibility of using automatically extracted routine clinical data in a respiratory cohort study: The SPHN–SPAC demonstrator project"

### SUPPLEMENTARY DISPLAY ITEMS

| Type of clinical event | Definition | Details |
|---|---|---|
| SPAC baseline visit | Outpatient clinical visit in pneumology outpatient clinic closest to the date of returned SPAC baseline questionnaire | Outpatient clinical event in pneumology outpatient clinic + diagnosis or lung function test on the same or closest date within the following 90 days |
| Pneumology follow up visit | Subsequent outpatient clinical visits in pneumology outpatient clinic | |
| Respiratory-related emergency department visits | Outpatient clinical visits in the emergency department for respiratory reasons | Outpatient clinical event in emergency department + free-text diagnosis with ≥ 1 respiratory keyword [1] |
| Respiratory-related hospitalizations | Inpatient clinical events with respiratory primary diagnosis | Inpatient clinical event with primary ICD-10 diagnosis = J00–J99, R05 or R06 |

**Abbreviations:** ICD-10 = International Classification of Diseases, 10th revision
[1] In supplementary box 2

**Supplementary Table 1: Definition of respiratory clinical events for SPHN-derived data**

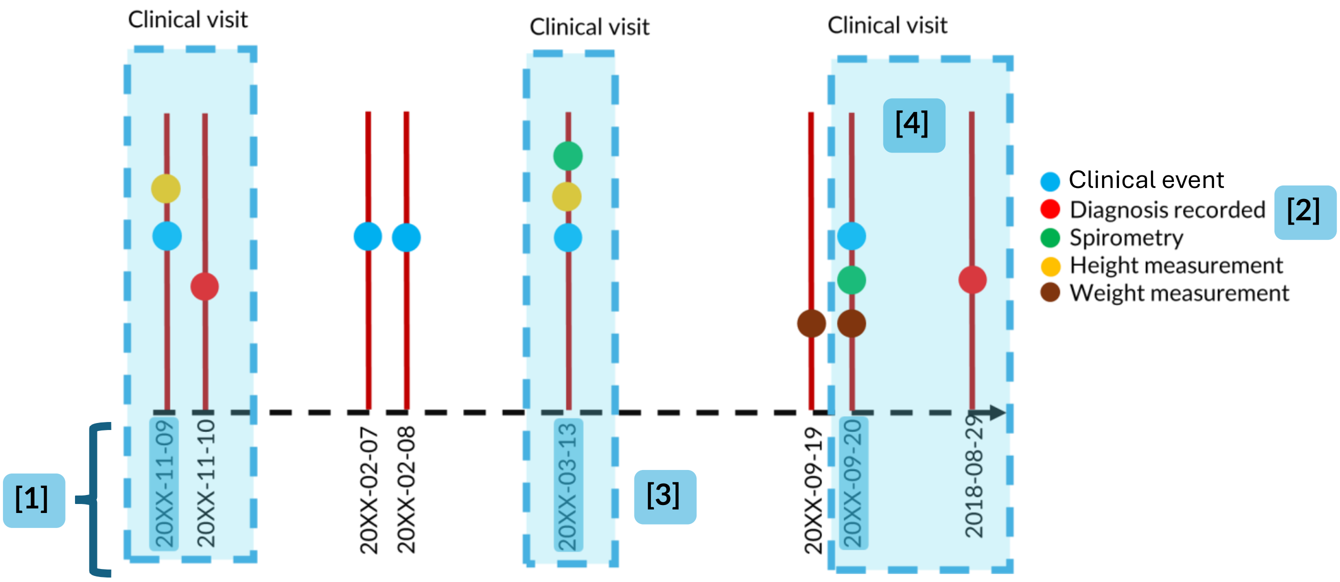


Because SPHN-derived outpatient data linkage relies on timestamped clinical events rather than explicit visit identifiers, we applied a multi-step procedure to reconstruct visit-centered outpatient clinical visits.

- **[1]** We Identified pneumology-related outpatient clinical events using SPHN concept metadata indicating care setting. These events did not necessarily correspond to distinct clinical visits and could represent planned consultations, isolated diagnostic procedures, or administrative documentation.
- **[Light blue box]** We allowed delayed documentation. Based on consultation with clinicians, we allowed for a lag of up to 90 days between the clinical encounter and documentation in the electronic health record.
- [2] We used free-text diagnoses as anchors to identify outpatient clinical visits. We assumed that each diagnosis indicated a clinical encounter if it could plausibly be linked to a corresponding clinical event.
- **[3]** We assigned clinical visit dates using the timestamp of the linked clinical event rather than the diagnosis documentation date.
- **[4]** We classified a clinical event as an outpatient clinical visit if a diagnosis could be linked to the clinical event within a 90-day window or if a lung-function test was recorded on the same calendar date as the clinical event. When both a diagnosis and a lung-function test were linked to the same event, they were treated as a single outpatient clinical visit.
    - We removed duplicate diagnoses within the same 90-day window by retaining the diagnosis with the smallest temporal distance to the corresponding clinical event. Diagnoses that could not be plausibly linked to any event were excluded.
- We assigned additional clinical variables (e.g. anthropometric measurements and laboratory results) to reconstructed visits based on temporal proximity, linking each variable to the most plausible outpatient visit.

**Supplementary Figure 1. Restructuring of SPHN-derived data into visit-centered clinical events**

**Supplementary table 2. Completeness of clinical variables for respiratory-related emergency department visits and hospitalizations from SPHN-derived data**

| Clinical variables | Hospital A n = 517 | Hospital B n =558 |
|---|---|---|
| **Respiratory related emergency department visit, n (%)** | | |
| Children, n | 53 | 83 |
| Number of events, n | 70 | 133 |
| Height from clinical records | 0 (0) | 22 (17) |
| Weight from clinical records | 69 (99) | 133 (100) |
| Diagnostic information | 70 (100) | 133 (100) |
| Treatment information | 18 (26) | 3 (2) |
| Spirometry | 0 (0) | 0 (0) |
| FeNO | 0 (0) | Not available |
| **Respiratory related hospitalization, n (%)** | | |
| Children, n | 18 | 20 |
| Number of events, n | 23 | 22 |
| Height from clinical records | 16 (70) | 12 (55) |
| Weight from clinical records | 21 (91) | 0 (0) |
| Diagnostic information | 23 (100) * | 22 (100) * |
| Treatment information | 18 (78) | 20 (91) |
| Spirometry | 0 (0) | 0 (0) |
| FeNO | NA | Not available |

**Abbreviations:** FeNO = fractional exhaled nitric oxide; ICD-10 = International Classification of Diseases, 10th revision

*Diagnosis information included primary in ICD 10 format

**Supplementary box 1: Acquisition and preparation of SPHN-derived data in the SPHN-SPAC demonstrator project**

SPHN-derived hospital data were obtained and prepared for analysis through an iterative, multi-step process conducted in close collaboration with clinicians, hospital IT teams, and the SPHN Data Coordination Centre (DCC):

**Identification of extractable variables**

Eligible SPAC variables were identified through joint review of the SPHN concept catalogue and local EHR documentation practices. Only variables represented by approved SPHN clinical concepts were considered extractable.

**Mapping to SPHN standards and data request**

Hospital IT teams mapped local EHR fields to SPHN terminology standards and populated predefined extraction tables. Formal data requests specified variables, coding rules, de-identification procedures, and extraction windows and required approval by participating hospitals and the SPHN DCC.

**Generation and transfer of RDF data**

Extraction tables were uploaded to the SPHN Connector, where they were converted into Resource Description Framework (RDF) format using international terminologies (ICD-10-GM, SNOMED CT, LOINC). RDF files were securely transferred to the BioMedIT platform.

**Transformation into visit-centered datasets**

Within BioMedIT, RDF files were converted to tabular format and reshaped into visit-centered datasets. Diagnoses, measurements, medications, and lung-function results were linked to the nearest plausible clinical encounter using a predefined 90-day window.

**Iterative validation and harmonization**

Transformed datasets were iteratively validated for completeness and coding consistency. Revised extracts were requested as needed. Validated encounters were harmonized into four predefined clinical event types and linked with manually abstracted SPAC data and self-reported healthcare utilization using pseudonymized identifiers.

**Abbreviations: ICD-10-GM, International Classification of Diseases, 10th Revision, German Modification; SNOMED CT, Systematized Nomenclature of Medicine—Clinical Terms; LOINC, Logical Observation Identifiers Names and Codes.**

#### Supplementary Methods: Identification of respiratory-related healthcare use

Respiratory-related healthcare events were identified using free-text diagnostic keywords for outpatient and emergency department (ED) visits and primary ICD-10 codes for hospitalizations. For ED visits, identification involved two steps: first, a broad keyword set was used to identify all respiratory-related records, including upper-respiratory conditions; second, a narrower keyword set was applied to identify visits likely to have been reported in response to the SPAC follow-up question about ED visits due to “cough, wheezing, breathing problems, or asthma.”

##### Broad extraction of respiratory-related clinical events

The initial extraction was performed by the hospital IT teams and included respiratory-related outpatient visits, ED visits, and hospitalizations. The core asthma- and wheeze-related keywords were informed by diagnoses recorded in the manually abstracted SPAC data and previous SPAC work on diagnostic labels for childhood asthma and wheeze.[1] Additional respiratory terms were included to broaden the extraction, and the list was refined iteratively with hospital clinicians, hospital IT staff, and the SPAC study team during data-quality checks. This broad extraction included both upper- and lower-respiratory conditions.

**Initial data extraction:** Outpatient and ED events were included if the available diagnostic text contained at least one predefined keyword. Hospitalizations were identified separately using primary ICD-10 diagnoses.

Before keyword matching, diagnostic text was converted to lowercase, and leading and trailing spaces were removed. The following criteria were applied to outpatient and ED records:

- **Substring matching:** bronch; pneumo; asthma; pleura; respir; strepto; laryng; atmung; husten; obstruktiv; pharyn; atemweg; luftweg; wheeze; rhinitis; airway; dyspnoe.
- **Approximate matching with a Levenshtein distance of ≤2 characters:** pseudokrupp; krupp-syndrom; grippaler; asthma; sinusitis; dyspnoe; rhinitis; wheeze; influenza; airway.
- **Exact matching:** krupp; grippe.

Substring matching identified terms appearing within longer words, such as “bronch” in “bronchitis,” while approximate matching allowed up to two-character differences to capture spelling variations or typographical errors.

##### Respiratory-related ED visits

Among ED visits identified in the broad extraction, we applied a narrower keyword set to identify visits likely to have been reported in the SPAC follow-up questionnaire as being due to “cough, wheezing, breathing problems, or asthma.” Terms referring only to upper-respiratory conditions were not included in this narrower list.

The ED-specific keywords were applied to the first 80 characters of the diagnostic text to reduce matches based on secondary diagnoses or past medical history.

The following criteria were applied:

- **Substring matching:** bronch; asthma; respir; pneumo; laryng; atmung; husten; obstruktiv; atemweg; luftweg; wheeze; airway; dyspnoe.

- **Approximate matching with a Levenshtein distance of ≤2 characters:** pseudokrupp; krupp-syndrom; asthma; dyspnoe; wheeze; influenza; airway; giemen; atemnot.
- **Exact matching:** krupp.

An ED visit was classified as respiratory-related for comparison with the questionnaire if at least one of these criteria was met.

**Manual assessment of potential misclassification**

To assess potential false negatives, we manually reviewed all ED visits included in the initial hospital extraction that had diagnostic text available but were not classified as respiratory-related by the ED keyword criteria. This included 21 visits in Hospital A and 62 visits in Hospital B. Only one visit, in Hospital B, was considered consistent with the SPAC follow-up question and was reclassified as a respiratory-related ED visit. The other 82 visits were considered correctly excluded by the ED criteria.

To assess potential false positives, we manually reviewed a random sample of 100 ED visits classified as respiratory-related, including 50 from each hospital. One visit was considered clearly unrelated because the respiratory diagnosis was documented only as part of the patient's past medical history and was therefore excluded from the final classification.

Residual false-positive classifications may remain when a respiratory condition was recorded as a secondary or previous diagnosis although the main reason for the ED visit was unrelated. Residual false-negative classifications may remain when relevant diagnoses were absent from the available text, documented using terminology not included in the keyword lists, or unavailable in the SPHN-derived data.

**Respiratory-related hospitalizations**

Hospitalizations were classified as respiratory-related when the primary diagnosis was coded within ICD-10 J00–J99, R05, or R06, including their subcodes. No narrower filter was applied because the identified primary diagnoses corresponded to the respiratory conditions covered by the SPAC follow-up question.

1. de Jong CCM, Pedersen ESL, Mozun R, Müller-Suter D, Jochmann A, Singer F, et al. Diagnosis of asthma in children: findings from the Swiss Paediatric Airway Cohort. *Eur Respir J.* 2020;56(5):2000132. doi:10.1183/13993003.00132-2020.

| Hospital A | Hospital B |
|---|---|
| 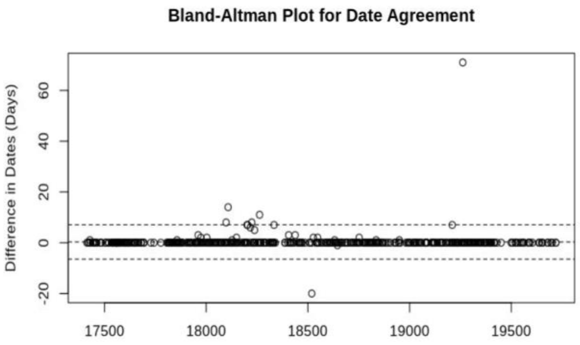 | 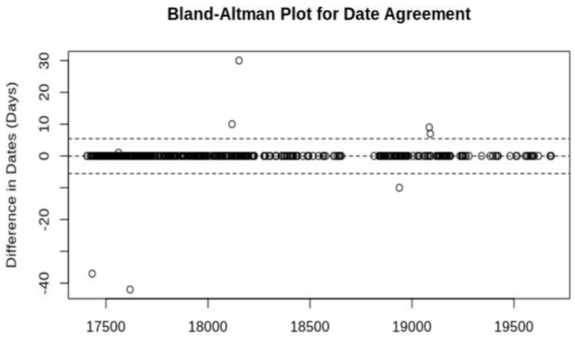 |





| | |
|---|---|
| *Difference in days (median, range) = 0 (0 - 71)* | *Difference in days (median, range) = 0 (0 - 53)* |
| *Exact matches (n, %) = 489 (94.6%)* | *Exact matches (n, %) = 550 (98.5%)* |

**Supplementary figure 2 Concordance of clinical visit date between manually abstracted clinical data and SPHN-derived hospital data for SPAC baseline visit. Bland-Altman plots**

| Hospital A | | | |
|---|---|---|---|
| **SPHN- derived data** / **Manually abstracted clinical data** | Female | Male | Total |
| Female | 210 | 3 | 213 |
| Male | 8 | 296 | 304 |
| Total | 218 | 299 | 517 |

*Cohen kappa (CI) = 0.96 (0.93 – 0.98)*

| Hospital B | | | |
|---|---|---|---|
| **SPHN- derived data** / **Manually abstracted clinical data** | Female | Male | Total |
| Female | 202 | 6 | 208 |
| Male | 7 | 343 | 350 |
| Total | 209 | 349 | 558 |

*Cohen kappa (CI) = 0.95 (0.92 – 0.98)*

**Supplementary figure 3. Concordance of sex (Cohen' kappa) between manually abstracted clinical data and SPHN-derived hospital data for SPAC baseline visit**

#### Concordance of anthropometric data Hospital A

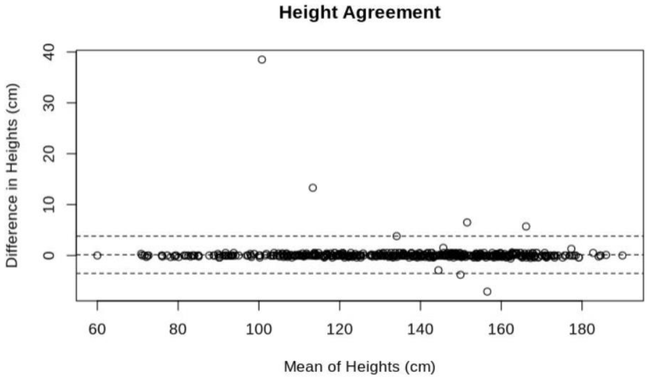


Concordance correlation coefficient: 0.96 (0.96–0.97)
n =318

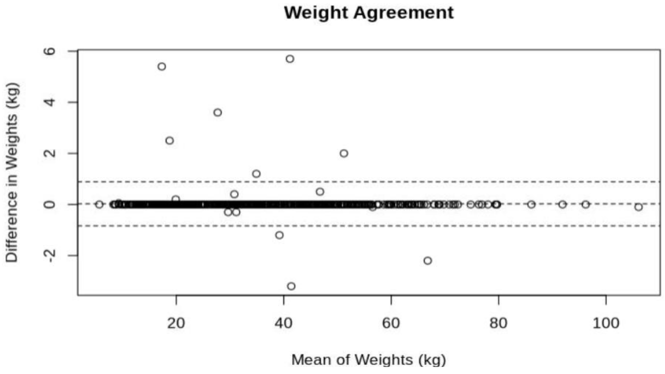


Concordance correlation coefficient: 0.92 (0.89–0.93)
n =321

**Demming regression height (cm)**

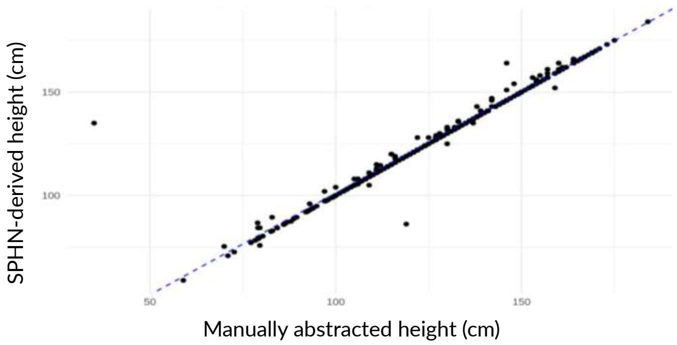


Demming regression

| | Coefficient | SD | |
|---|---|---|---|
| Intercept | 2.89 | 3.09 | -3.17 – 8.95 |
| Slope | 0.982 | 0.02 | 0.93 –2.025 |

**Demming regression weight (kg)**

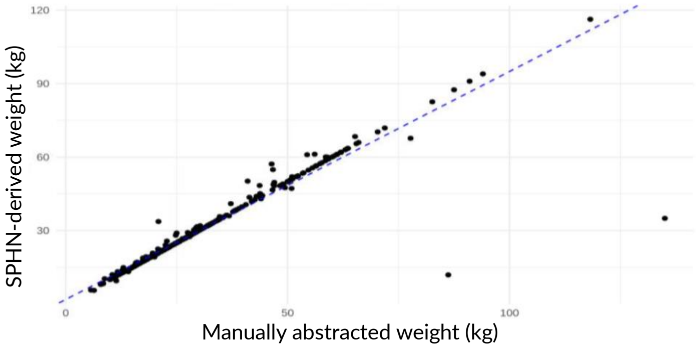


Demming regression

| | Coefficient | SD | CI |
|---|---|---|---|
| Intercept | 1.86 | 1.48 | -1.05 – 4.76 |
| Slope | 0.931 | 0.06 | 0.813 – 1.05 |

**Supplementary figure 4. Concordance of anthropometric measures from medical records (height and weight) between manually abstracted clinical data and SPHN-derived data for SPAC baseline visit for <u>Hospital A</u>**

#### Concordance of anthropometric data Hospital B

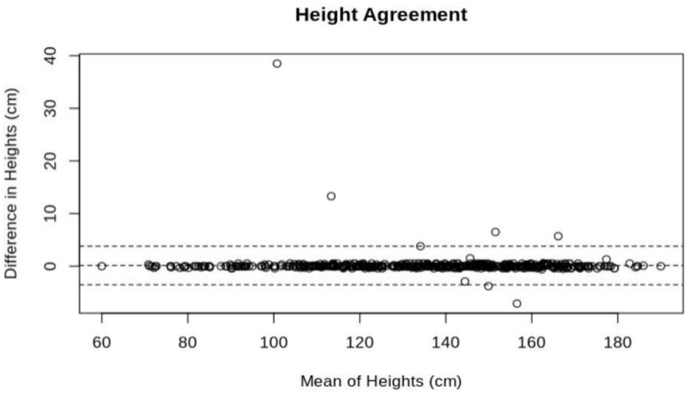


Concordance correlation coefficient: 0.997 (0.996 - 0.998) n =529

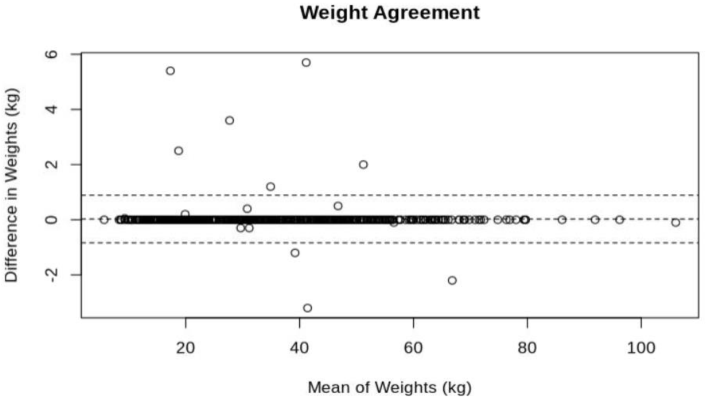


Concordance correlation coefficient: 0.999 (0.999 - 0.999) n =532

**Demming regression height (cm)**

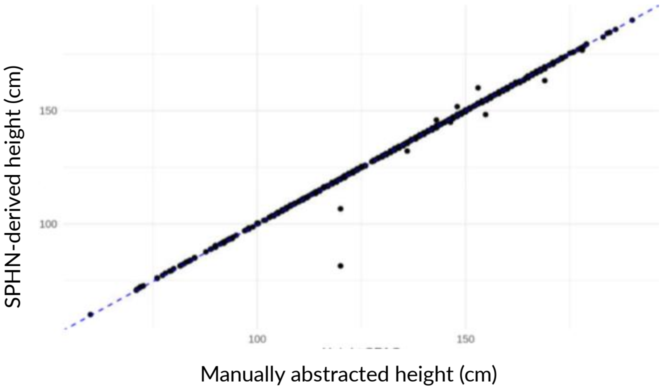


Demming regression

| | Coefficient | SD | CI |
|---|---|---|---|
| Intercept | 0.666 | 0.583 | -1.8 – 0.047 |
| Slope | 1.003 | 0.003 | 0.996 –1.011 |

**Demming regression weight (kg)**

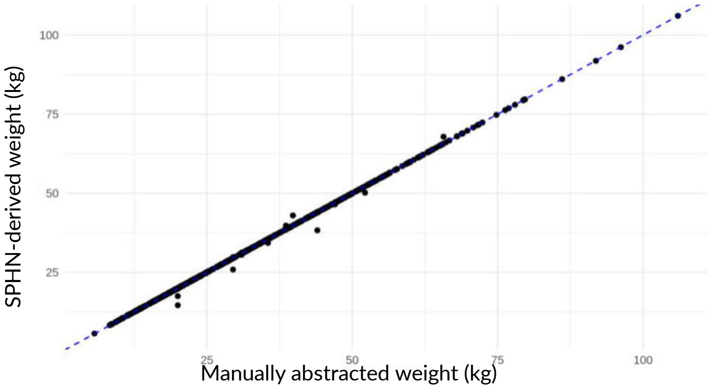


Demming regression

| | Coefficient | SD | CI |
|---|---|---|---|
| Intercept | -0.070 | 0.038 | -0.147 – 0.005 |
| Slope | 1.001 | 0.001 | 0.999 –1.003 |

**Supplementary figure 5. Concordance of anthropometric measures from medical records (height and weight) between manually abstracted clinical data and SPHN-derived data for SPAC baseline visit for Hospital B.**

**Hospital A**

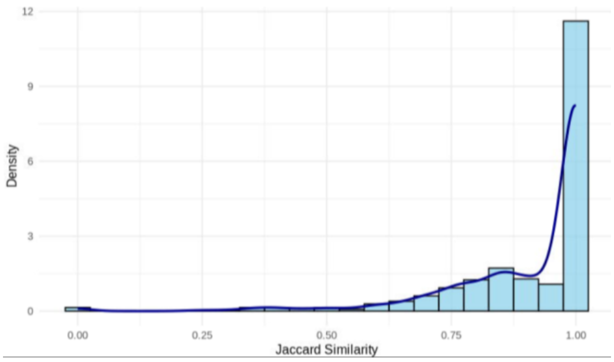


Exact matches (Jaccard =1), n (%) =309 (60%)
Jaccard similarity, mean (IQR) = 0,92 (0.91-1.00)
Same diagnostic information (Jaccard >0.4) = 505/517

**Hospital B**

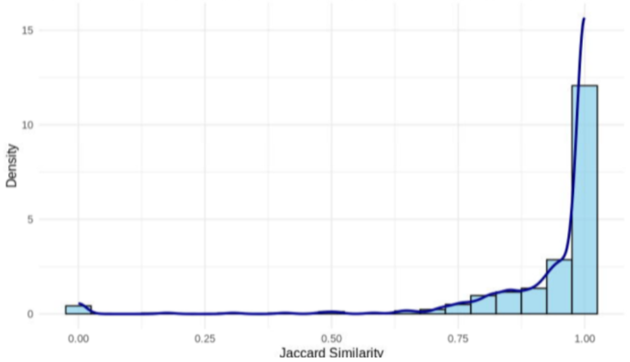


Exact matches (Jaccard =1), n (%) =323 (58%)
Jaccard similarity, mean (IQR) = 0,90 (0.84-1.00)
Same diagnostic information (Jaccard >0.3) = 552/558

**Supplementary figure 6. Concordance of free-text diagnosis (distribution of Jaccard similarity index scores) between manually abstracted clinical data and SPHN-derived data for SPAC baseline visit for Hospital A and B.**

#### Concordance of spirometry values (FEV1 and FVC) Hospital A

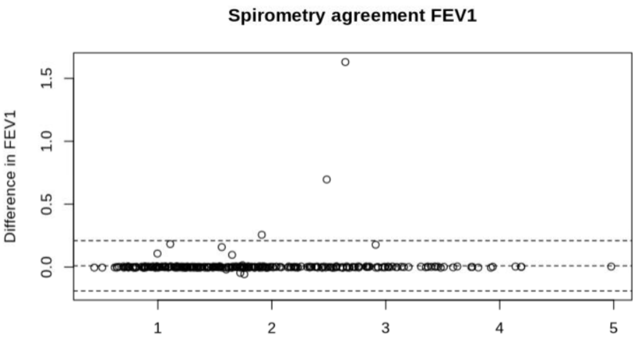


Concordance correlation coefficient: 0.992 (0.990 - 0.994) n =318

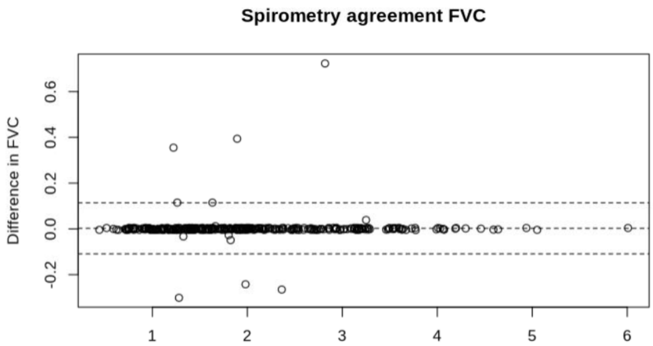


Concordance correlation coefficient: 0.998 (0.998 - 0.998) n =327

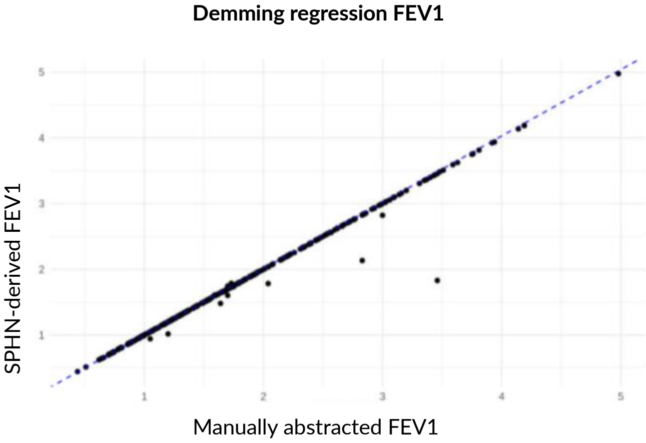


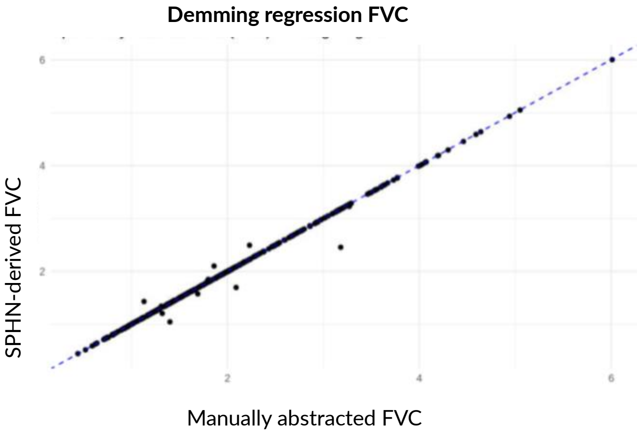


Demming regression

| | Coefficient | SD | CI |
|---|---|---|---|
| Intercept | -0.005 | 0.007 | -0.020 – 0.009 |
| Slope | 1.009 | 0.007 | 0.995 –1.022 |

Demming regression

| | Coefficient | SD | CI |
|---|---|---|---|
| Intercept | -0.002 | 0.005 | -0.009 – 0.009 |
| Slope | 1.001 | 0.002 | 0.997 –1.006 |

**Supplementary figure 7. Concordance of spirometry values (FEV1 and FVC) between manually abstracted clinical data and SPHN-derived data for SPAC baseline visit for Hospital A.**

#### Concordance of spirometry values (FEV1 and FVC) Hospital B

Spirometry agreement FEV1

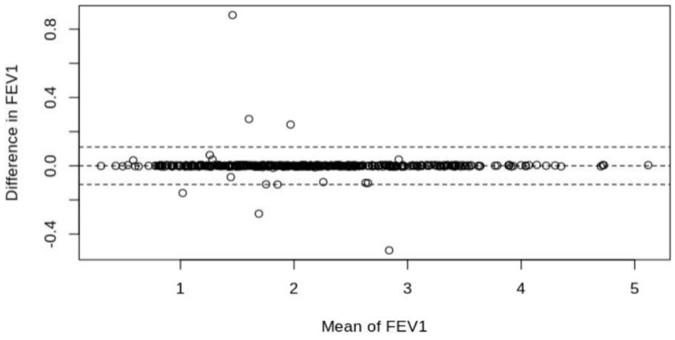


Concordance correlation coefficient: 0.997 (0.997–0.998)
n =426

Spirometry agreement FVC

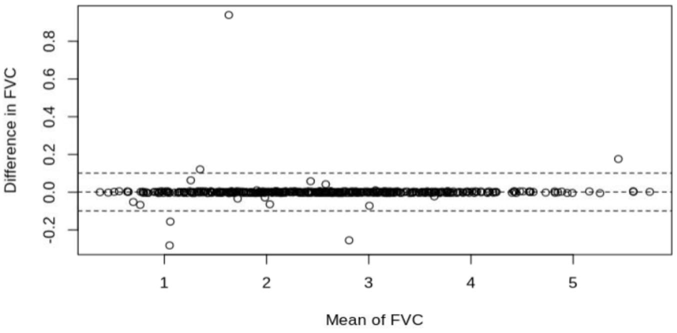


Concordance correlation coefficient: 0.998 (0.998–0.999)
n =432

**Demming regression FEV1**

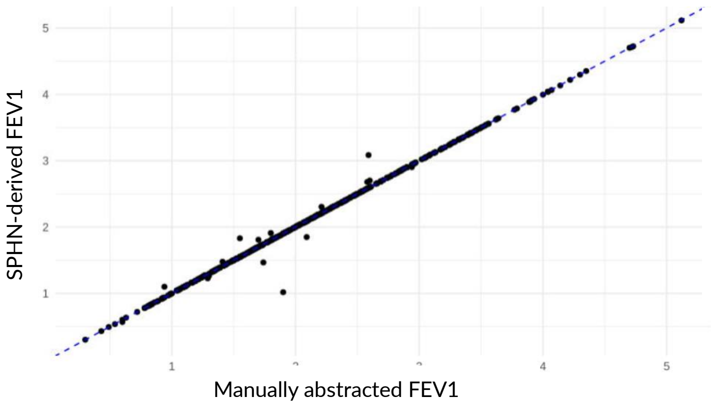


Demming regression

| | Coefficient | SD | CI |
|---|---|---|---|
| Intercept | 0.006 | 0.007 | -0.008– 0.020 |
| Slope | 0.997 | 0.002 | 0.992– 1.002 |

**Demming regression FVC**

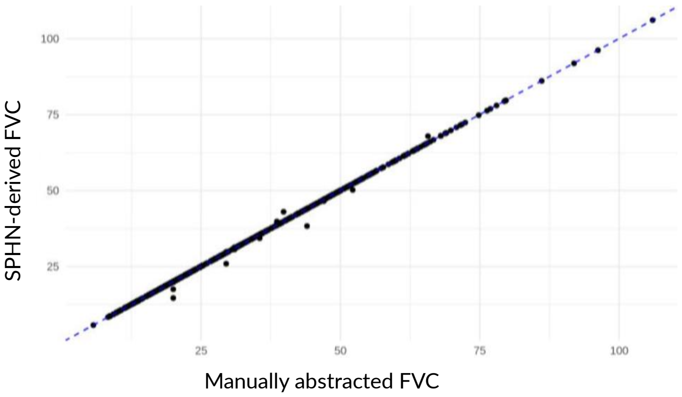


Demming regression

| | Coefficient | SD | CI |
|---|---|---|---|
| Intercept | 0.000 | 0.008 | -0.016– 0.015 |
| Slope | 1.000 | 0.003 | 0.996– 1.005 |

**Supplementary figure 8. Concordance of spirometry values (FEV1 and FVC) between manually abstracted clinical data and SPHN-derived data for SPAC baseline visit for Hospital B.**
